# An extensible laboratory information management system for data harmonization across research centers: The ICTS Dashboard

**DOI:** 10.64898/2026.05.31.26354439

**Authors:** Charles Hadley King, Ivan De Dios, Rebekah Barrik, Seth Berger, Miguel Almalvez, Lightning Auriga, Emmanuèle Délot, Changrui Xiao, Jonathan LoTempio, Eric Vilain

## Abstract

**Background:** Collaborative research programs increasingly require infrastructure capable of integrating heterogeneous participant, sample, and experimental data while meeting evolving research needs. Existing tools, including clinical EHRs, REDCap, generic research information management systems, and bespoke database builds, were not designed to operationalize externally governed, versioned consortium data models across database structure, APIs, validation, web forms, and reproducible exports. The ICTS Dashboard, developed at the University of California, Irvine Institute for Clinical and Translational Science, fills this need by providing a general purpose research information management system.

**Methods:** The ICTS Dashboard was built as an open-source, schema-driven platform in which database structure, server-side validation, REST APIs, web-based forms, and reproducible exports are derived from a versioned JavaScript Object Notation (JSON) Schema set. The backend is implemented in Django, Django REST Framework, and PostgreSQL; the frontend in React. We instantiate the platform with the Genomics Research to Elucidate the Genetics of Rare Diseases (GREGoR) Data Model and extend it with two case studies: a locally developed biobank table for biospecimen logistics, and an embedded adaptation of the RAG-HPO retrieval-augmented phenotype curation tool.

**Results:** The ICTS Dashboard deployed at the UCI-GREGoR site supports 37 schema-derived tables and 250 documented API endpoints. It holds metadata for 2,563 participants, 1,238 families, 5,517 biobank entries, 2,593 sequenced experiments, and 289 genetic findings. It supports quarterly external data submissions regenerated directly from the database. The biobank extension adds entities the consortium does not standardize while preserving foreign-key linkage to rare disease records; the RAG-HPO module adds curator-mediated phenotype normalization against 19,389 indexed HPO terms. Both were integrated without modifying the GREGoR Data Model.

**Conclusion:** The Dashboard’s architecture is not limited to GREGoR or to rare disease research. A version-controlled, machine-readable data model can serve not only as a data sharing standard but as the operational backbone of a research program when paired with schema-governed tooling. Any collaborative research program with a structured, versioned model can adopt the same pattern to reduce implementation overhead and improve reproducibility, harmonization, and findable, accessible, interoperable, and reusable (FAIR)-aligned data management.

## Introduction

Large-scale genomics programs increasingly require the integration of heterogeneous participant data, including phenotypic descriptions, biospecimen catalogues, genomic and multi-omic files, and tertiary genomic analysis from case review [1,2]. Investigators frequently depend on institution-level electronic health record systems (EHRs), such as Epic, and research data capture systems, such as REDCap [3–6]. While REDCap is highly customizable for study-specific data capture, adapting it to support complex and evolving data models can require substantial effort, technical expertise, and ongoing maintenance.

These systems provide robust support for clinical operations or study data collection, but were not designed as interoperable tools for consortium-scale management of harmonized participant, biospecimen, phenotype, and genomic research data. Existing systems operate within fragmented institutional environments where information is distributed across disparate databases and applications, limiting interoperability and creating barriers to efficient data integration and reuse [7]. Prior studies have demonstrated that semantic harmonization and standardized data models are necessary to preserve meaning across systems and to support downstream applications such as dashboards, decision support tools, and research workflows [7,8].

These limitations are especially relevant for large, multi-center rare disease genomics initiatives such as the Genomics Research to Elucidate the Genetics of Rare Diseases (GREGoR) Consortium [1]. To support this work across five research centers, the GREGoR Consortium Data Coordinating Center (DCC) led a working group effort to create a version-controlled data model for harmonizing participant, phenotype, family, biospecimen, sequencing, and analysis data across sites.

That group produced the GREGoR Data Model [9], which standardizes an analysis-ready consortium dataset through regular public data releases. Participating centers submit validated quarterly data releases to the NHGRI Genomic Data Science Analysis, Visualization, and Informatics Lab-space (AnVIL), a federated cloud platform for genomic data sharing and analysis [10].

Here, we describe the design and implementation of the ICTS Dashboard, a data model-agnostic tool to facilitate research collaborations (Fig 1). We instantiate the dashboard with the GREGoR Data Model, with focus on how version-controlled GREGoR table definitions can be converted into JSON Schemas that govern database structures, validation rules, APIs, and web-based forms, allowing local data management to remain aligned with evolving consortium releases.

**Figure 1:**
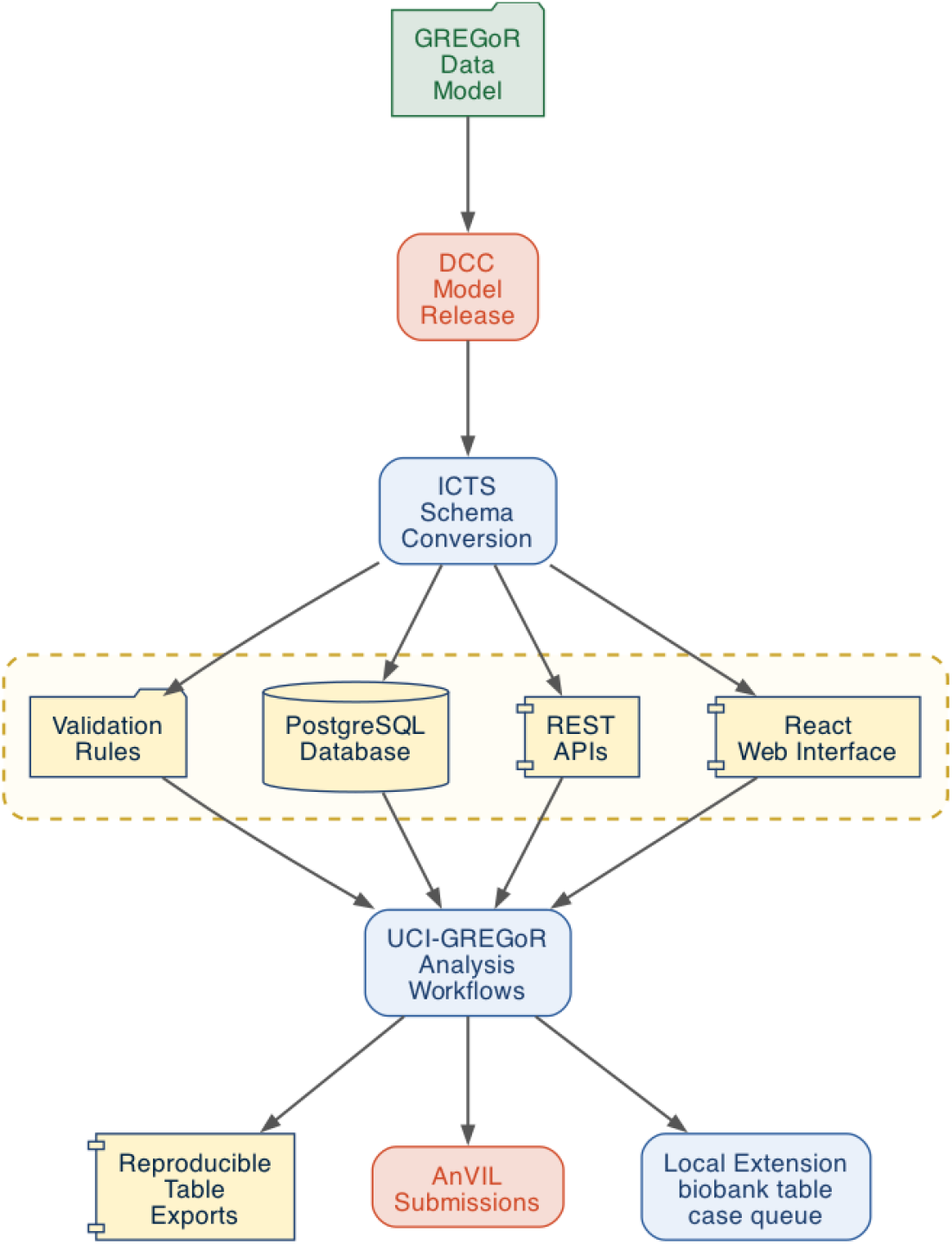
Conceptual workflow for developing the ICTS Dashboard from the GREGoR Data Model. The version-controlled GREGoR Data Model is released as machine-readable table definitions, which are converted into JSON Schemas for ICTS Dashboard implementation. These schemas govern database structures, validation rules, application programming interfaces, web-based forms, and export workflows. The same schema-governed architecture supports UCI-GREGoR operational workflows, reproducible exports, AnVIL submissions, and local extensions such as the biobank table

Using the UCI-GREGoR deployment as a case study, we show how this approach supports harmonized tracking of participants, phenotypes, biospecimens, sequencing data, and genetic findings; reproducible AnVIL submissions; and integration with external research systems. More broadly, this work demonstrates how to enhance the usability of consortium data models to better function as reusable, open-source infrastructure that other genomics programs can adapt for translational research workflows.

## Materials and Methods

### Schema processing workflow

The ICTS Dashboard was designed as a platform for processing JSON schemas. The Dashboard was designed with the GREGoR Data Model as the first use case [9]. The GREGoR DCC instantiates the model as a series of linked tables that can be viewed as spreadsheets and serialized into machine-readable formats, including JSON, LinkML, and DBML. For the ICTS Dashboard, we used the JSON representation released through the GREGoR DCC repository [11].

The tables were organized within the Dashboard into metadata and experiment categories. Metadata tables store descriptive information used to identify related participants and interpret genetic findings. Experiment tables provide provenance for multi-omic sequencing and analysis workflows, documenting the experimental data used to generate variant calls and support genetic findings. While based on the GREGoR Data Model as a use case, the Dashboard’s JSON schema processing engine is project agnostic.

The ICTS Dashboard uses the same versioned GREGoR JSON Schemas across data entry, validation, storage, and export. Records entered or updated through the Dashboard are validated against the active schema version before being saved and later exported. This allows generation of database records that have been structured according to the corresponding GREGoR Data Model release.

Because each export is tied to a defined schema version, exports can be regenerated from the same database-backed source with consistent field definitions, required values, and table structures. This workflow reduces manual reformatting, limits divergence between local records and consortium submission tables, and supports reproducible exports.

Schema and export-related code are maintained under version control, allowing changes to validation behavior or export structure to be reviewed alongside updates to the supported GREGoR Data Model release.

### ICTS Dashboard Platform Architecture

The ICTS Dashboard backend was implemented using Django[12], Django REST Framework[13], and PostgreSQL[14]. GREGoR-derived JSON Schemas were used to guide table implementation and schema-level server-side validation. This enables each supported GREGoR table to be represented as a Django model, or structured database table. API access is authenticated using JSON Web Tokens (JWT)[15], and endpoints are documented through OpenAPI/Swagger[16] using “drf-yasg”[17].

For each data model table, the backend exposes the standard create, read, update, and delete operations to support the entry, review, and correction of data. Delete operations are restricted to administrator accounts assigned by the Django ‘is_superuser’ flag. We also implemented a few broader, read-only custom endpoints for retrieval of aggregate data across multiple tables. This creates a separation of table-level data management and cross-table data queries used for reporting and export.

ICTS Dashboard database models are linked through foreign key relationships to preserve dependencies among participant, family, biospecimen, phenotype, sequencing, and findings records. Deletion behavior was intentionally constrained to prevent accidental loss of downstream records. For example, participant deletion requires associated metadata and experiment records to be reviewed or removed in dependency order rather than relying on automatic cascading deletion.

The ICTS Dashboard frontend was implemented in ReactJS [18] and implements an additional validation layer for user-facing and context-specific checks before API submission. These checks include immediate field-level feedback for required fields, type mismatches, restricted enumerations, identifier formatting, table-specific input normalization, and selected conditional requirements that are difficult to represent in Draft-07 JSON Schema or impractical to enforce uniformly at the API layer.

The active GREGoR schema version is defined in the application environment configuration, allowing each deployment to load the schema set corresponding to a specific GREGoR Data Model release. The compiled application uses this configured version to load the corresponding JSON Schema files and generate table views, data entry forms, and edit interfaces for each supported GREGoR table.

Schema properties are parsed to determine field names, expected data types, required fields, controlled vocabularies (enumerations), and multi-value inputs. Field descriptions and help text embedded in the schemas are displayed as help text within the user interface, allowing contributors to reference GREGoR field definitions during data entry.

Bulk imports are supported through both the web interface and backend API. On the frontend, a table uploader page allows users to select a target table and schema, upload a tab-separated values (TSV) file, and convert each row into a JSON object according to the selected schema. The resulting list of objects is submitted to the backend API for validation and creation. On the backend, bulk upload requests include metadata identifying the target table and a list of JSON objects to be validated and inserted using the corresponding table-specific logic.

Role-based permissions were used to reduce the risk of accidental data loss and restrict access to sensitive operations. There are two roles in the dashboard: standard users, and administrator users. Standard users can create, view, and update records according to their assigned permissions. Administrative users can flag records for review, hiding those records from standard users or take destructive actions, including record deletion.

Model relationships are configured to prevent automatic cascading deletion or invalidation of related entries, requiring dependent participant, phenotype, biospecimen, sequencing, and findings records to be reviewed before parent records are removed.

The ICTS Dashboard was designed to manage research metadata separately from protected health information (PHI) and personally identifiable information (PII). Identifiable participant information is maintained in HIPAA-compliant systems such as REDCap, while the Dashboard is designed to store GREGoR-aligned metadata. This separation allows research workflows to be coordinated through the Dashboard while limiting unnecessary exposure of identifiable participant data.

The ICTS Dashboard uses continuous integration (CI) with Django’s built-in testing tool over a set of testing data [12]. Every GitHub pull request runs through CI testing before it can be merged with the main branch. The demo data is designed to simulate API requests for every table and various frontend-backend interactions that occur in supported workflows under the design constraints. The testing data is designed to validate the enforcement of constraints implemented at the model and database query level.

### Integration with external tooling

RAG-HPO is an open-source Python-based tool that uses retrieval-augmented generation (RAG) to extract phenotype phrases and support normalization to Human Phenotype Ontology (HPO) terms[19]. Developed by BCM-GREGoR Center[20], the original RAG-HPO repository provided the retrieval-augmented approach for extracting phenotype phrases, retrieving candidate HPO terms from embedded HPO text, and then querying a large language model to support term selection[20,21].

Our implementation extended this workflow by embedding it within the ICTS Dashboard as a persistent, reviewable phenotype curation module designed to assist genetic counselors and clinicians during data entry. This adaptation preserves manual review, provenance, and compatibility with the GREGoR Data Model phenotype table. The Django HPO app implements models for HPO terms, ontology edges, and versioned vector artifacts. The PostgreSQL table provides full-text search and autocompletion. The DRF APIs expose the HPO module to the React frontend. The React-based interface allows users to inspect candidate terms, replace selected terms, export results, and import reviewed HPO annotations into the Dashboard phenotype table.

### Open-source Software Availability

The ICTS Dashboard is available as open-source software through the UCI-ICTS GitHub Organization (https://github.com/UCI-ICTS/icts-dashboard/releases/). The repository includes the Django backend, React frontend, schema conversion utilities, deployment documentation, and testing data used for testing and documentation. Releases are versioned and archived to preserve the relationship between source code, generated JSON schemas, and validation/export behavior.

## Results

### Dashboard instantiation

The ICTS Dashboard is instantiated for UCI-GREGoR using the GREGoR Data Model for the underlying structure. At the time of writing, it supports 37 GREGoR schemas/tables and contains metadata from 2,563 participants, 1,238 families, 5,517 biospecimens, 2,593 sequenced experiments, and 289 genetic findings. The results to follow describe several implemented components that demonstrate the utility of this platform for use with other structured research data models.

### Schema-based APIs and UI

The version-controlled GREGoR Data Model is converted into JSON Schemas, and those are used to define the database structure, validation rules, application programming interfaces (APIs), and web-based forms. The ICTS Dashboard exposes 250 API endpoints (Table 1) across metadata, experiment, authentication, search, HPO, S3, administrative, and documentation services (https://genomics.icts.uci.edu/api/redoc/).

**Table 1.**
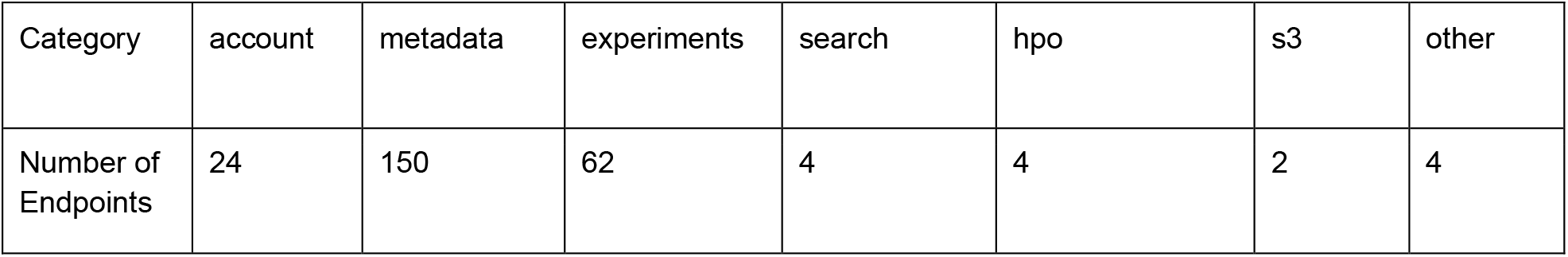
API endpoints exposed by the ICTS Dashboard. Endpoints are grouped by application domain and reflect the current API surface of the ICTS Dashboard. Metadata endpoints support participant, family, phenotype, biospecimen, prior testing, and analyte-related records. Experiment endpoints support sequencing, analysis, variant provenance, and genetic findings workflows. Additional endpoint groups support account and user management, custom search, HPO/RAG-HPO functionality, secure S3-based access to sequence data resources. The server health, administrative utilities and automatically generated API documentation through Swagger and ReDoc, each of which is a single endpoint, are all combined in “other”.

Most endpoints support metadata management (150 endpoints) and experiment data management (62 endpoints), reflecting the scope of the GREGoR Data Model and the need to manage both descriptive research metadata and sequencing-derived evidence. Metadata endpoints support records such as participant, family, phenotype, biospecimen, prior testing, and analyte data, while experiment endpoints support sequencing workflows, variant provenance, and genetic findings. This API structure allows the Dashboard to function as an operational layer over the GREGoR Data Model, supporting table-specific data entry, cross-table views, quality-control workflows, and reproducible exports.

Frontend table entry forms are generated from the active schema version, with the field descriptions from the schema becoming the help text in the data entry forms. This schema-derived interface provides immediate feedback on each field for invalid or missing values, including errors related to required fields, type mismatches, and restricted enumerations.

We also implemented selected cross-table specific validations that are difficult to express or impractical to enforce through the backend via JSON schema validation alone, such as context-dependent field requirements, identifier formatting, and table-specific input normalization. One such example is how the genetic findings table implements conditional requirements with Django selector methods based on the GREGoR JSON Schema description.

Backend API validation provides the authoritative schema-level gate for persisted records, while frontend validation improves usability by highlighting affected fields before submission and applying selected context-specific checks. This layered approach allowed the Dashboard to support both programmatic API submission and guided web-based data entry.

### Validation and reproducible exports

The Dashboard supports bulk data import via a web-based table uploader and direct API bulk submission. Bulk imports allow users to upload table-formatted records into the Dashboard while applying schema-based validation before records are accepted, helping identify missing required fields, invalid data types, restricted enumeration values, and table-specific formatting issues early in the workflow. The table uploader extended schema-based validation to bulk data entry. Users can select a target table and schema, upload a TSV file, and submit the converted JSON records through the API. This allowed large batches of records to be imported using the same validation and table-specific logic applied to individual create requests.

Once imported or entered through the web interface, records are stored in the database according to the active GREGoR Data Model release. This allows exports to be regenerated from a consistent database-backed source rather than manually assembled from independent spreadsheets. Because validation rules, export logic, and schema versions are maintained under version control, the same records can be repeatedly exported with consistent field definitions, required values, and table structures. This workflow supports external data deposition while reducing manual reformatting and limiting divergence between local operational records and consortium-formatted data releases.

### Semi-automated generation

Early ICTS Dashboard database models were implemented manually from GREGoR-derived schemas. To reduce repetitive development effort, we developed a semi-automated workflow that converts spreadsheet-based table specifications into JSON Schemas and then uses those schemas to generate initial Django model structures. Generated models still require manual review and refinement before production use, but they provide a consistent starting point for database implementation. In contrast, Dashboard web forms are generated directly from the active schema definitions, allowing new or revised tables to inherit field definitions, validation rules, controlled vocabularies, and help text without requiring custom form development. This significantly reduced the need for custom development for new tables, while preserving manual review for the backend.

### Extensibility of the dashboard

The core GREGoR Data Model supports biospecimen, analyte, sequencing, and analysis records, but it does not explicitly capture local biobank operations such as freezer location, box position, shipment tracking from clinical laboratories to sequencing centers, or sample processing status prior to downstream data generation. At UCI-GREGoR we were utilizing a biospecimen management workflow that was composed of multiple spreadsheets, with each sheet created to track a different portion of the specimen processing workflow. To streamline this workflow we developed a local biobank table that links operational biospecimen tracking to GREGoR participant records.

The biobank extension captures required identifiers and status fields, including biobank_id, participant_id, received_date, specimen_type, and status, while also supporting freezer, shelf, rack, box, tube barcode, shipment date, tracking number, requested test, and downstream derivative analyte fields. Status values track the local sample lifecycle from pending shipment through storage, extraction, data delivery, variant analysis readiness, QC issues, loss, or replacement requests. This table therefore extends the Dashboard beyond consortium submission requirements by supporting day-to-day biospecimen logistics while preserving links to GREGoR participant, analyte, experiment, and alignment records.

The biobank table demonstrates how the ICTS Dashboard can incorporate site-specific workflow needs without changing the core GREGoR Data Model. In this case, the extension allowed local sample handling, freezer tracking, shipment management, and downstream analysis readiness to be managed within the same Dashboard environment used for GREGoR participant, phenotype, sequencing, and findings workflows.

The ICTS Dashboard incorporated RAG-HPO as a phenotype curation module compatible with the GREGoR phenotype table. The active HPO artifact was generated from the Human Phenotype Ontology OBO source file on 2026-02-17. It includes 19,389 HPO terms, 23,677 parent-child ontology edges, and 24,209 synonym entries across 11,028 terms. PostgreSQL search fields were populated for 19,388 terms, supporting autocomplete, full-text search, and fuzzy spelling lookup. The derived HPO text artifact contained 19,389 rows and was linked to a FAISS vector index containing 19,389 text-embedding-3-large embeddings with 3,072 dimensions.

RAG-HPO results are displayed in an interactive review table where each extracted phenotype phrase is paired with a selected HPO term and an expandable ranked candidate set. Genetic counselors and clinicians can inspect candidates, replace selected terms, download results, or import reviewed annotations into the Dashboard phenotype table after assigning a participant_id. Manual replacements are retained as user-selected choices, keeping the workflow curator-mediated rather than fully automated.

## Discussion

The Dashboard usefully fills a collaborative data sharing niche that is not well served by currently available commercial or open source platforms. REDCap remains excellent for IRB-bound participant intake, consent, and survey data, and we continue to rely on it for exactly those purposes; it is not designed, however, to manage biospecimen logistics, sequencing provenance, and consortium-formatted export against an externally versioned data model. Epic and similar EHRs are clinical systems and do not expose the structured research metadata that consortia work or general research may require.

General-purpose research information management systems can offer flexible project administration but do not target genomics-specific entities such as analytes, alignments, and genetic findings, nor can they adapt their schemas to consortia standards. Custom Django builds against a data model are common at individual sites, but they typically encode the model implicitly in handwritten models and serializers, which is precisely the drift problem the Dashboard is built to eliminate.

The Dashboard is therefore most useful when three conditions for a research program hold: First, the program has a versioned, machine-readable data model. Next, the program has intake needs that exceed what that model captures. And finally, the program needs to meet reproducible data sharing obligations.

As expectations for broad sharing of human genomic data continue to increase[22], research programs need infrastructure that can support local operations while producing consistent, reusable data releases. The ICTS Dashboard addresses this gap by connecting schema-based data entry, validation, database storage, and reproducible exports within a single workflow.

### Extensibility as proof of concept for generalizability

While the GREGoR data model serves as the backbone trial case examined in this paper, we extend that model through the addition of a biobank schema and external tooling (RAG-HPO). The biobank table extends the model horizontally: it adds entities (freezer position, shipment tracking, processing status) that the consortium does not standardize but that local laboratory operations require, while preserving a foreign-key relationship to GREGoR participant records. RAG-HPO extends the model vertically: it does not add new tables, but augments an existing one (phenotype) with a curator-mediated workflow for converting free-text clinical descriptions into ontology-aligned terms.

Both were absorbed without forking the upstream GREGoR model; the biobank table reused schema-derived interfaces, while RAG-HPO required a dedicated review interface. Iterative refinement of the biobank specification, driven by user feedback through GitHub issues, converted several free-text fields into enumerations, reducing spelling variation and consolidating tracking that had previously lived in parallel spreadsheets.

### Limitations and the cost of schema-driven design

The Dashboard has not yet been deployed against a non-GREGoR data model, so the generalizability claim, while structurally supported, remains to be demonstrated in another context. Semi-automated generation of Django models from JSON Schemas reduces, but does not eliminate, manual implementation, and generated models still require review before production use.

The schema-driven approach also carries costs that should be acknowledged. When the upstream model changes between releases, the database, validators, and export logic must all be reconciled against the new schema; for additive changes this is straightforward, but renamed fields, retyped columns, or restructured enumerations require coordinated migration. Given the Dashboard’s flexibility over multiple new versions of the GREGoR data model, we think this limitation may paradoxically support its generalizability to other data models.

Some validation rules cannot be expressed cleanly in Draft-07 JSON Schema or enforced uniformly through table-level API validation. The clearest example is the GREGoR genetic findings table, where assigning a “solved” status depends on a checklist of criteria spanning multiple fields and conditional dependencies.

These checks are currently implemented in the React form layer before API submission. As a result, the backend enforces schema-level validity, while the frontend enforces additional workflow-specific validity for entering data through the user interface.

We treat this as a deliberate stratification of validation. Schema-level checks are enforced at the API layer, while context-dependent checks are concentrated at the point of data entry where field-level guidance is the most useful. Unifying this logic across the frontend and backend remains a future development goal.

RAG-HPO must remain curator-mediated - specifically, a member of our case review team manually approves the addition of new information from this large language model-supported tool. Source phenotype descriptions imported from REDCap may contain PHI or PII that is clinically meaningful but not appropriate for transmission to embedding services, and sanitization is currently a manual step.

Adoption by other groups will require technical expertise. Documentation and GitHub issue tracking help, but migration from existing workflows is non-trivial; the biobank transition at UCI required several months of parallel operation before users were confident the system stored and returned the fields they expected.

## Future directions

Four priorities follow from the limitations above. First, we plan to deploy the Dashboard against a non-GREGoR data model. The ICTS supports the UCI Center for Rare Diseases which includes an Undiagnosed Diseases Network site and the Differences of Sex Development Translational Research Network, both of which maintain distinct data structures [23,24]. Instantiating the Dashboard against these will validate the generalizability claim directly. Second, we will integrate the dashboard with our IRB-approved scripted chatbot Kauro[25] for integration of data on enrollment as a future schema. Third, we intend to push more validation logic (particularly conditional requirements) out of React and the JSON Schemas and into the Django generation layer itself, taking advantage of features beyond Draft-07 where appropriate. This would simplify maintenance and make backend and frontend generation of forms, APIs, and database structures more automated. Fourth, we will extend automated testing to the frontend, where coordinated schema-form behavior currently relies on user verification.

### Conclusions

The ICTS Dashboard shows that a version-controlled, machine-readable data model can serve not only as a submission standard but as the operational backbone of a translational genomics program. By treating schemas as the primary source of truth for database structure, APIs, validation, user interfaces, and export, the platform collapses what are typically separate engineering efforts into one governed workflow. The GREGoR deployment, the locally developed biobank extension, and the embedded RAG-HPO module together demonstrate that this architecture accommodates consortium standards, site-specific operations, and community tooling without forking the underlying model.

Although built around GREGoR, the Dashboard’s design is not intrinsic to that project or even rare disease. Any consortium or research program that maintains a structured, versioned data model could adopt the same pattern to reduce implementation overhead and improve reproducibility, harmonization, and FAIR-aligned accessibility. We offer the platform as open-source infrastructure to support that broader reuse.

## Data Availability

The ICTS Dashboard source code, schema conversion utilities, documentation, and release materials are available through the UCI-ICTS GitHub repository at https://github.com/UCI-ICTS/icts-dashboard/releases/. The GREGoR Data Model used to generate the Dashboard schemas is available through the GREGoR Data Coordinating Center repository(https://github.com/UW-GAC/gregor_data_models/releases). Aggregate implementation metrics reported in this manuscript, including schema/table counts, API endpoint counts, and HPO artifact summary statistics, are included in the manuscript. Participant-level data managed within the ICTS Dashboard are not publicly available because they contain sensitive research and clinical metadata governed by institutional review board protocols, data use agreements, and participant consent. GREGoR consortium data releases are made available through NHGRI AnVIL according to GREGoR and AnVIL access policies.

https://github.com/UCI-ICTS/icts-dashboard

